# Nirmatrelvir and the Risk of Post-Acute Sequelae of COVID-19

**DOI:** 10.1101/2022.11.03.22281783

**Authors:** Yan Xie, Taeyoung Choi, Ziyad Al-Aly

## Abstract

Long Covid – the disease encompassing the post-acute sequelae of SARS-CoV-2 (PASC) —affects millions of people around the world. Prevention of PASC is an urgent public health priority. In this work, we aimed to examine whether treatment with nirmatrelvir in the acute phase of COVID-19 is associated with reduced risk of post-acute sequelae. We used the healthcare databases of the US Department of Veterans Affairs to identify users of the health system who had a SARS-CoV-2 positive test between March 01, 2022 and June 30, 2022, were not hospitalized on the day of the positive test, had at least 1 risk factor for progression to severe COVID-19 illness and survived the first 30 days after SARS-CoV-2 diagnosis. We identify those who were treated with oral nirmatrelvir within 5 days after the positive test (n=9217) and those who received no COVID-19 antiviral or antibody treatment during the acute phase of SARS-CoV-2 infection (control group, n= 47,123). Inverse probability weighted survival models were used to estimate the effect of nirmatrelvir (versus control) on a prespecified panel of 12 post-acute COVID-19 outcomes and reported as hazard ratio (HR) and absolute risk reduction (ARR) in percentage at 90 days. Compared to the control group, treatment with nirmatrelvir was associated with reduced risk of PASC (HR 0.74 95% CI (0.69, 0.81), ARR 2.32 (1.73, 2.91)) including reduced risk of 10 of 12 post-acute sequelae in the cardiovascular system (dysrhythmia and ischemic heart disease), coagulation and hematologic disorders (deep vein thrombosis, and pulmonary embolism), fatigue, liver disease, acute kidney disease, muscle pain, neurocognitive impairment, and shortness of breath. Nirmatrelvir was also associated with reduced risk of post-acute death (HR 0.52 (0.35, 0.77), ARR 0.28 (0.14, 0.41)), and post-acute hospitalization (HR 0.70 (0.61, 0.80), ARR 1.09 (0.72, 1.46)). Nirmatrelvir was associated with reduced risk of PASC in people who were unvaccinated, vaccinated, and boosted, and in people with primary SARS-CoV-2 infection and reinfection. In sum, our results show that in people with SARS-CoV-2 infection who had at least 1 risk factor for progression to severe COVID-19 illness, treatment with nirmatrelvir within 5 days of a positive SARS-CoV-2 test was associated with reduced risk of PASC regardless of vaccination status and history of prior infection. The totality of findings suggests that treatment with nirmatrelvir during the acute phase of COVID-19 reduces the risk of post-acute adverse health outcomes.

## Introduction

Long Covid – the disease encompassing the post-acute sequelae of SARS-CoV-2 infection — affects millions of people around the world^1,2^. Despite this substantial toll, there is no approved medication for the prevention or treatment of Long Covid. Several hypotheses have been proposed to explain the underlying mechanisms of Long Covid including persistence of the virus (or its fragments) or intensity of the inflammation during the acute phase of the disease^3^. The antiviral nirmatrelvir (in combination with ritonavir) which has been shown to reduce the risk of progression to severe acute COVID-19 has been suggested as a candidate drug that may reduce the risk of developing Long Covid^4,5^. In December 2021, oral nirmatrelvir was approved in the US for the treatment of acute SARS-CoV-2 infection (typically within 5 days of symptom onset) in non-hospitalized people at risk of progression to severe COVID-19 illness. Millions of people in the US have since received treatment with nirmatrelvir. Urgent calls have been made to evaluate whether treatment with nirmatrelvir in the acute phase of COVID-19 reduces the risk of Long Covid – but data has thus far been lacking^4^. Addressing this question will guide treatment approaches of SARS-CoV-2 infections and will inform the effort to develop and optimize prevention and treatment strategies for Long Covid.

In this analysis, we use the healthcare databases of the US Department of Veterans Affairs to identify users of VA health system who had a SARS-CoV-2 positive test between March 01, 2022 and June 30, 2022, were not hospitalized on the day of the positive test, had at least 1 risk factor for progression to severe COVID-19 illness, and survived the first 30 days after SARS-CoV-2 diagnosis; we identify those who were treated with oral nirmatrelvir within 5 days after the positive test and did not receive other outpatient COVID-19 antiviral or antibody treatment within 30 days after the positive test (nirmatrelvir group, n=9217) and those who received no outpatient COVID-19 antiviral or antibody treatment within 30 days after the positive test (control group, n= 47,123). We then used inverse probability weighting approach to balance the characteristics of the groups and evaluate whether treatment with oral nirmatrelvir versus control is associated with reduced risk of post-acute outcomes including post-acute sequelae of COVID-19 (PASC) defined as having at least one post-acute sequela (from a set of 12 prespecified post-acute sequelae of SARS-CoV-2 infection), post-acute death, post-acute hospitalization and each individual post-acute sequela.

## Methods

### Setting

The VA operates the largest integrated health care system in the US; the system is comprised of 1,293 healthcare facilities (including 171 VA Medical Centers and 1,112 outpatient sites) located across the US. The VA provides comprehensive healthcare to discharged veterans of the US armed forces including preventative and health maintenance, outpatient care, inpatient hospital care, prescriptions, mental healthcare, home healthcare, primary care, specialty care, geriatric and extended care, medical equipment, and prosthetics.

### Data sources

The study was conducted using the healthcare databases of the US Department of Veterans Affairs. VA healthcare data are updated daily. VA healthcare databases include individual-level demographic information and data on healthcare encounters, comorbidities, procedures, and surgeries. Data domains included outpatient encounters, inpatient encounters, inpatient and outpatient medications, and laboratory results. The VA COVID-19 Shared Data Resource was used to collect information on COVID-19 patients and vaccination status. The Area Deprivation index (ADI) — which is a composite measure of income, education, employment, and housing — was used as summary measure of contextual disadvantage at participants’ residential locations^6^.

### Cohort

A flowchart and a timeline of cohort construction are provided in supplemental figure 1 and 2, respectively. There were 67,579 participants who had a positive SARS-CoV-2 test result between March 01, 2022 and June 30, 2022, where their first date of positive test was set to be T_0_. Because nirmatrelvir is an outpatient treatment for COVID-19, we further selected 62,549 participants who were not hospitalized at T_0;_ 9424 participants treated with nirmatrelvir within 5 days of T_0_ were selected into the nirmatrelvir group. We then selected 9353 participants with at least one risk factor of progression to severe acute COVID-19 illness, which included age>60, BMI>25 km/m^2^, current smoker, cancer, cardiovascular disease, kidney disease, chronic lung disease, diabetes, immune dysfunction and hypertension. We also removed participants with liver disease, end stage kidney disease or eGFR<30 mL/min/1.73m^2^ (n=9248). Participants who did not use other outpatient COVID-19 antiviral or antibody treatments within 30 days after T_0_ were selected (n=9239). To examine the post-acute events, only participants alive 30 days after T_0_ were included in the nirmatrelvir group (final n=9217).

A control group of participants was constructed from the 62,549 participants who had a positive SARS-CoV-2 test result between March 01, 2022 and June 30, 2022 and did not hospitalized at the date of positive test (T_0_). Within those who did not receive nirmatrelvir within 5 days of T_0_ (n=53,125), we selected 52,352 participants with at least one risk factor of progression to severe acute COVID-19 illness, and removed participants with liver disease, end stage kidney disease or eGFR<30 mL/min/1.73m^2^ (n=49,661). Participants who did not use any outpatient COVID-19 antiviral or antibody treatments within 30 days after T_0_ (n=47,534) were further selected. To examine the post-acute events, only participants alive 30 days after T_0_ were included in the control group (final n=47,123).

The final cohort consisted of 56,340 participants, of which 9217 were in nirmatrelvir group, and 47,123 in control group. The cohort was followed until August 31, 2022.

### Outcomes

Outcomes examined in this study included PASC – defined as having at least one sequela from a set of sequelae based on prior evidence^1,2,7-21^. We also examined the risk of post-acute death and hospitalization and a composite outcome of death or hospitalization. We also studied individual sequela including ischemic heart disease, dysrhythmia, deep vein thrombosis, pulmonary embolism, fatigue, liver disease, acute kidney injury, muscle pain, diabetes, neurocognitive impairment, shortness of breath and cough. Individual sequelae were defined based on inpatient and outpatient ICD-10 diagnosis codes and laboratory results; death was defined based on vital status data; hospitalization was defined based on inpatient encounter data; and the outcome of post-acute sequelae was defined at the time of first occurrence of any sequelae included in the study (individual sequela, death, or hospitalization). Incident outcomes were assessed within those without history of the related outcome within one year before T_0_. All outcomes were ascertained 30 days after T_0_.

### Covariates

We identified baseline characteristics that may influence the use of treatment and the occurrence or assessment of outcomes based on literature review and prior knowledge^1,14,20,22,23^. All covariates were assessed within three years before study enrollment unless otherwise specified. Predefined covariates included age, race (White, Black, and Other), sex, ADI, body mass index (BMI), smoking status (current, former, and never), prior history of SARS-CoV-2 infection, use of steroids, use of long-term care, eGFR, systolic and diastolic blood pressure, cancer, chronic lung disease, dementia, diabetes, hyperlipidemia and immune dysfunction. We also considered healthcare utilization parameters including number of outpatient and inpatient encounters, number of laboratory encounters and number of outpatient medications received within one year before study enrollment and influenza vaccination status. We additionally specified pandemic related characteristics including week of the SARS-CoV-2 positive test result, hospital bed capacity, hospital bed occupancy at the participants’ health care facility within the week of the SARS-CoV-2 positive test result. Continuous variables were transformed into restricted cubic spline functions to account for potential non-linear relationships.

### Statistical Analysis

Baseline characteristics were reported as mean and standard deviation or frequency and percentage. Covariate balance between groups was evaluated by the absolute standardized differences where an absolute standardized difference of less than 0.1 was considered evidence of good balance.

To examine the risk of incident outcomes, for each outcome besides death or hospitalization, we conducted analysis on a sub-cohort of participants without the history of the outcome within one year before T_0_. An inverse probability weighting method was used to balance the differences in baseline characteristics between the nirmatrelvir and control groups. Logistic regression was built to predict the probability of receiving nirmatrelvir given covariates. The probability was then used as the propensity score. We then constructed the inverse probability weights as a value of 1 for those in the nirmatrelvir group and as propensity score/(1-propensity score) for those in the control group. Weights larger than 10 would be truncated at 10 to reduce the influence of extreme weight (in our data, no weights were larger than 10 and none were truncated). The inverse probability weights were then applied to a Cox survival model in order to estimate the effect of nirmatrelvir. Risk on the relative scale was reported as the hazard ratio and risk on the absolute scale was reported as the absolute risk reduction of nirmatrelvir compared to the control group at 90 days based on the survival probability difference between nirmatrelvir and control group.

The effect of nirmatrelvir on the risk of PASC was further examined within prespecified subgroups by age (≤60, >60-≤70 and >70 years), race (White and Black), sex, smoking status (current smoker, former smoker and never smoker), cancer, cardiovascular disease, chronic kidney disease, chronic lung disease, diabetes, Immune dysfunction and hypertension. We also examined the effect within populations with different vaccination status (unvaccinated, 1-2 doses of vaccine and boosted) and infection status (with primary SARS-CoV2 infection and reinfection). To examine the effect of nirmatrelvir within populations with different baseline risks, we also defined subgroups based on the number of baseline risk factors (1-2, 3-4 or ≥5) of progression to severe acute COVID-19 illness, where risk factors included age>60, BMI>25 km/m^2^, current smoker, cancer, cardiovascular disease, kidney disease, chronic lung disease, diabetes, immune dysfunction and hypertension.

We challenged the robustness of findings in multiple sensitivity analyses including 1) application of the overlap weighting method to balance baseline characteristics in the treatment and control groups (whereas in the primary approach, we used the inverse probability weighing approach to balance the groups); 2) application of the doubly robust approach to additionally adjust for covariates in the inverse probability weighted survival models (whereas in the primary approach we used inverse probability weighted survival models); 3) application of the high dimensional variable selection algorithm to additionally identify 100 covariates from data domains including diagnoses, medications and laboratory test results that were used along with predefined variables to construct the weights (whereas in the primary approach we used only predefined covariates); 4) adjusted for health care utilization during follow up including outpatient encounters, hospitalizations, laboratory tests and medications as time varying covariates (whereas in the primary approach, we used covariates information at T_0_).

Analyses were performed with SAS Enterprise Guide, version 8.2 (SAS Institute, Cary, NC). Data visualizations were performed in R 4.0.4 (R Foundation for Statistical Computing, Vienna, Austria). The robust sandwich variance estimator was used to estimate variance in weighted analyses. Risk on relative scale with 95% CI that does not cross 1 and risk on absolute scale with 95% CI that does not cross 0 was considered statistically significant. The study was approved by the US Department of Veteran Affairs St. Louis Health Care System Institutional Review Board which granted a waiver of informed consent.

## Results

The cohort included 56,340 participants; 9217 were treated with nirmatrelvir and 47,123 received no COVID-19 antiviral or antibody treatment within the first 30 days after infection; the latter group served as the control group. The demographic and health characteristics before weighting are provided in supplemental table 1; characteristics after weighting are provided in table 1. Absolute standardized mean differences between the nirmatrelvir and the control group after application of inverse probability weighting were all below 0.1 – suggesting good balance (table 1).

**Table 1.**
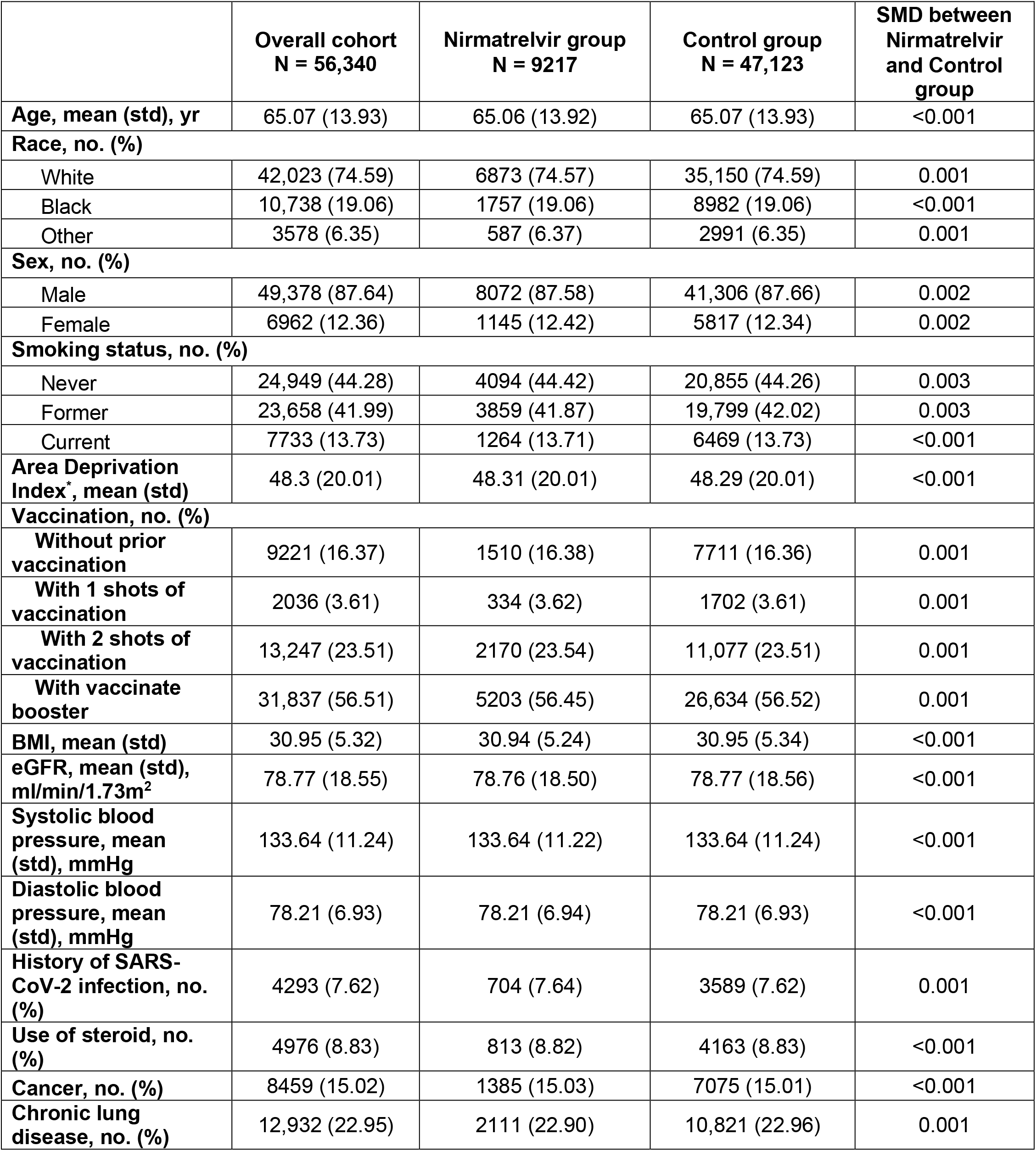
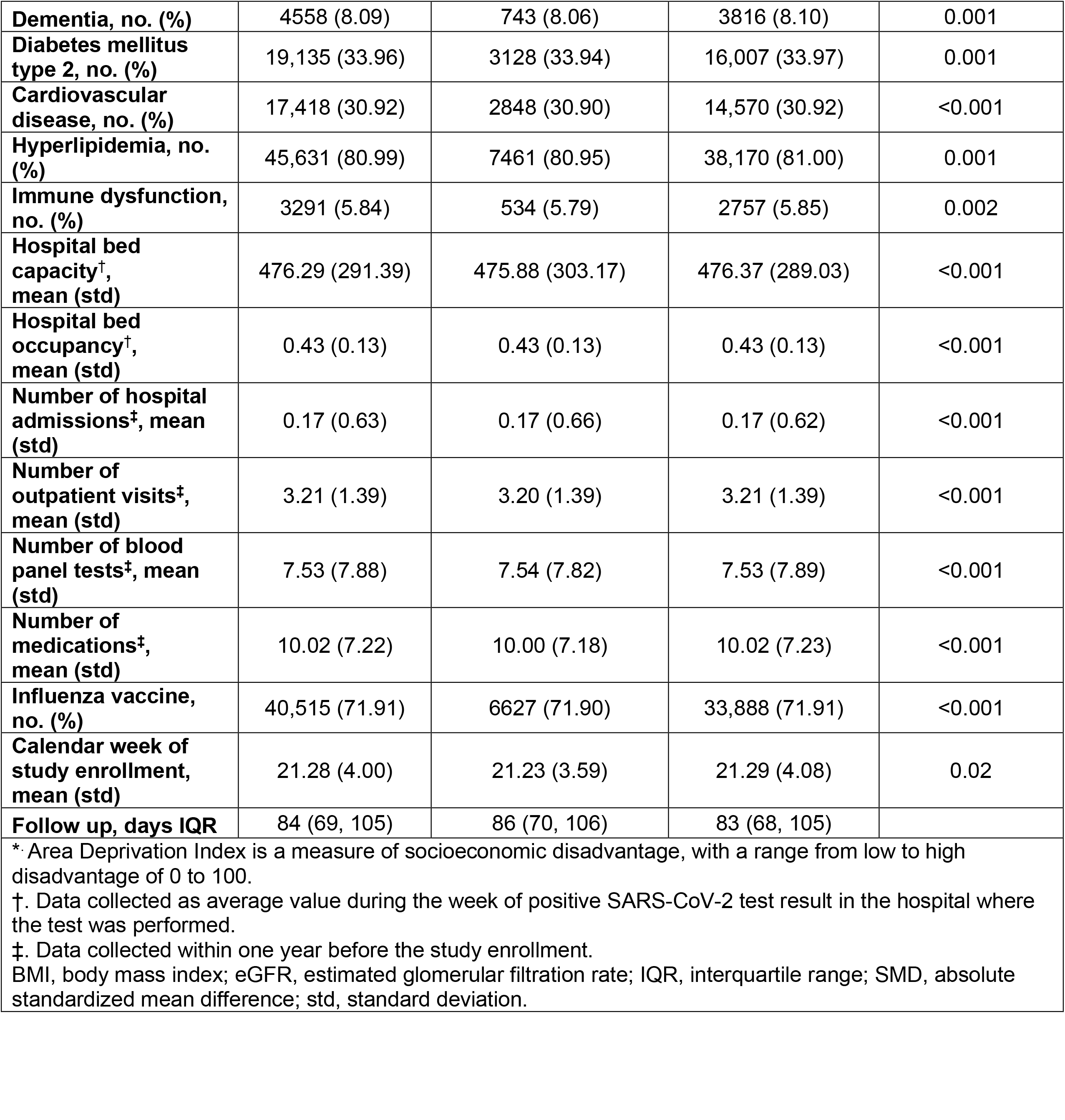
Demographic and health characteristics of the overall cohort, the nirmatrelvir group, and the control group after weighting.

|  | **Overall cohort**<br>**N = 56,340** | **Nirmatrelvir group**<br>**N = 9217** | **Control group**<br>**N = 47,123** | **SMD between**<br>**Nirmatrelvir**<br>**and Control**<br>**group** |
| --- | --- | --- | --- | --- |
| **Age, mean (std), yr** | 65.07 (13.93) | 65.06 (13.92) | 65.07 (13.93) | <0.001 |
| **Race, no. (%)** |  |  |  |  |
| White | 42,023 (74.59) | 6873 (74.57) | 35,150 (74.59) | 0.001 |
| Black | 10,738 (19.06) | 1757 (19.06) | 8982 (19.06) | <0.001 |
| Other | 3578 (6.35) | 587 (6.37) | 2991 (6.35) | 0.001 |
| **Sex, no. (%)** |  |  |  |  |
| Male | 49,378 (87.64) | 8072 (87.58) | 41,306 (87.66) | 0.002 |
| Female | 6962 (12.36) | 1145 (12.42) | 5817 (12.34) | 0.002 |
| **Smoking status, no. (%)** |  |  |  |  |
| Never | 24,949 (44.28) | 4094 (44.42) | 20,855 (44.26) | 0.003 |
| Former | 23,658 (41.99) | 3859 (41.87) | 19,799 (42.02) | 0.003 |
| Current | 7733 (13.73) | 1264 (13.71) | 6469 (13.73) | <0.001 |
| **Area Deprivation Index\*, mean (std)** | 48.3 (20.01) | 48.31 (20.01) | 48.29 (20.01) | <0.001 |
| **Vaccination, no. (%)** |  |  |  |  |
| Without prior vaccination | 9221 (16.37) | 1510 (16.38) | 7711 (16.36) | 0.001 |
| With 1 shots of vaccination | 2036 (3.61) | 334 (3.62) | 1702 (3.61) | 0.001 |
| With 2 shots of vaccination | 13,247 (23.51) | 2170 (23.54) | 11,077 (23.51) | 0.001 |
| With vaccinate booster | 31,837 (56.51) | 5203 (56.45) | 26,634 (56.52) | 0.001 |
| **BMI, mean (std)** | 30.95 (5.32) | 30.94 (5.24) | 30.95 (5.34) | <0.001 |
| **eGFR, mean (std), ml/min/1.73m<sup>2</sup>** | 78.77 (18.55) | 78.76 (18.50) | 78.77 (18.56) | <0.001 |
| **Systolic blood pressure, mean (std), mmHg** | 133.64 (11.24) | 133.64 (11.22) | 133.64 (11.24) | <0.001 |
| **Diastolic blood pressure, mean (std), mmHg** | 78.21 (6.93) | 78.21 (6.94) | 78.21 (6.93) | <0.001 |
| **History of SARS-CoV-2 infection, no. (%)** | 4293 (7.62) | 704 (7.64) | 3589 (7.62) | 0.001 |
| **Use of steroid, no. (%)** | 4976 (8.83) | 813 (8.82) | 4163 (8.83) | <0.001 |
| **Cancer, no. (%)** | 8459 (15.02) | 1385 (15.03) | 7075 (15.01) | <0.001 |
| **Chronic lung disease, no. (%)** | 12,932 (22.95) | 2111 (22.90) | 10,821 (22.96) | 0.001 |
| **Dementia, no. (%)** | 4558 (8.09) | 743 (8.06) | 3816 (8.10) | 0.001 |
| **Diabetes mellitus type 2, no. (%)** | 19,135 (33.96) | 3128 (33.94) | 16,007 (33.97) | 0.001 |
| **Cardiovascular disease, no. (%)** | 17,418 (30.92) | 2848 (30.90) | 14,570 (30.92) | <0.001 |
| **Hyperlipidemia, no. (%)** | 45,631 (80.99) | 7461 (80.95) | 38,170 (81.00) | 0.001 |
| **Immune dysfunction, no. (%)** | 3291 (5.84) | 534 (5.79) | 2757 (5.85) | 0.002 |
| **Hospital bed capacity<sup>†</sup>, mean (std)** | 476.29 (291.39) | 475.88 (303.17) | 476.37 (289.03) | <0.001 |
| **Hospital bed occupancy<sup>†</sup>, mean (std)** | 0.43 (0.13) | 0.43 (0.13) | 0.43 (0.13) | <0.001 |
| **Number of hospital admissions<sup>‡</sup>, mean (std)** | 0.17 (0.63) | 0.17 (0.66) | 0.17 (0.62) | <0.001 |
| **Number of outpatient visits<sup>‡</sup>, mean (std)** | 3.21 (1.39) | 3.20 (1.39) | 3.21 (1.39) | <0.001 |
| **Number of blood panel tests<sup>‡</sup>, mean (std)** | 7.53 (7.88) | 7.54 (7.82) | 7.53 (7.89) | <0.001 |
| **Number of medications<sup>‡</sup>, mean (std)** | 10.02 (7.22) | 10.00 (7.18) | 10.02 (7.23) | <0.001 |
| **Influenza vaccine, no. (%)** | 40,515 (71.91) | 6627 (71.90) | 33,888 (71.91) | <0.001 |
| **Calendar week of study enrollment, mean (std)** | 21.28 (4.00) | 21.23 (3.59) | 21.29 (4.08) | 0.02 |
| **Follow up, days IQR** | 84 (69, 105) | 86 (70, 106) | 83 (68, 105) |  |

\*. Area Deprivation Index is a measure of socioeconomic disadvantage, with a range from low to high disadvantage of 0 to 100.
†. Data collected as average value during the week of positive SARS-CoV-2 test result in the hospital where the test was performed.
‡. Data collected within one year before the study enrollment.
BMI, body mass index; eGFR, estimated glomerular filtration rate; IQR, interquartile range; SMD, absolute standardized mean difference; std, standard deviation.

In this study, we provide 2 estimates of risk: a) hazard ratio (HR) of nirmatrelvir in comparison to the control group and b) absolute risk reduction (ARR) in percentage at 90 days; the latter represents the event rate reduction in nirmatrelvir group compared to the control group at 90 days.

### Risk of PASC

We examined the risk of PASC defined as having at least one post-acute sequela (from a set of 12 prespecified post-acute sequelae of SARS-CoV-2 infection). Compared to the control group, nirmatrelvir was associated with reduced risk of PASC (HR 0.74 (95% CI 0.69, 0.81); the event rate was 9.43 (95% CI 9.14, 9.72) and 7.11 (95% CI 6.57, 7.64) per 100 persons at 90 days in the control and the nirmatrelvir groups, respectively. This corresponded to an ARR of 2.32 (95% CI 1.73, 2.91)) per 100 persons at 90 days (Figure 1 and 2a, supplemental table 2).

**Figure 1:**
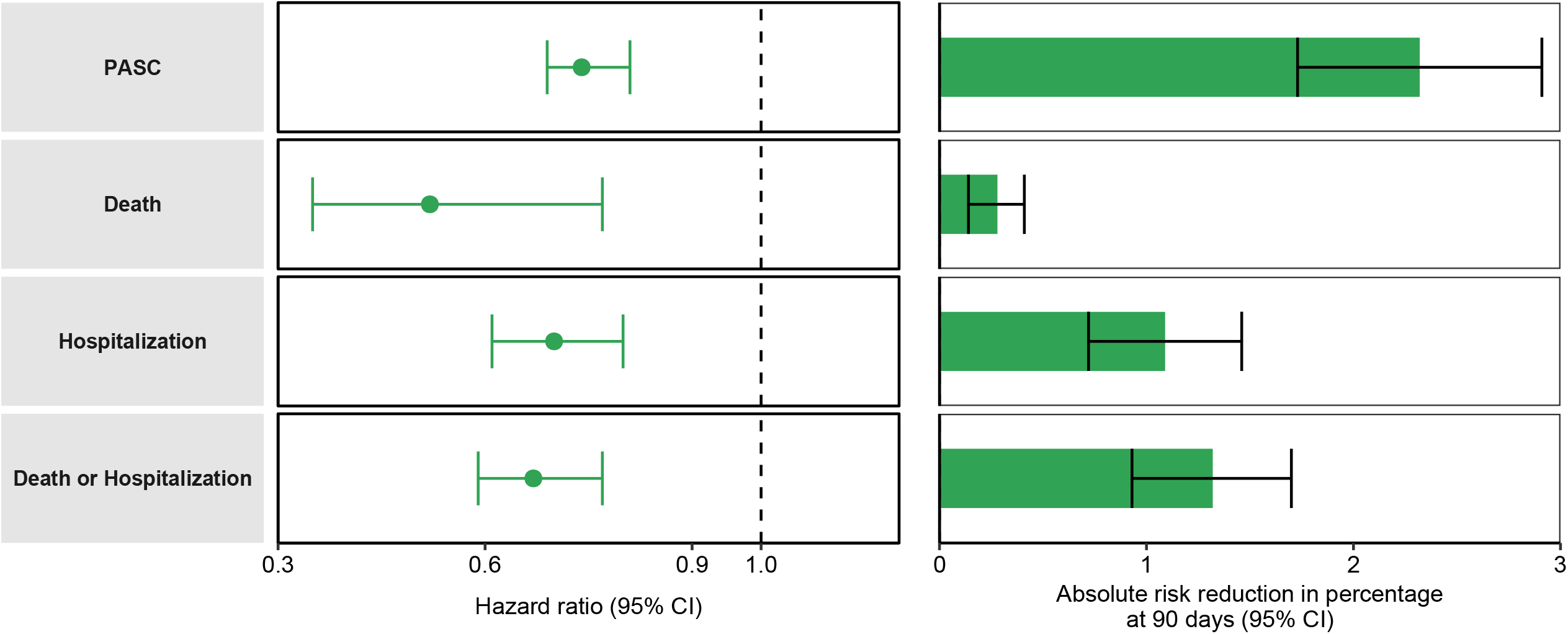
Hazard ratio and absolute risk reduction of nirmatrelvir on post-acute sequelae, death, hospitalization, and composite outcome of death or hospitalization compared to no treatment control group. Outcomes were ascertained 30 days after the SARS-CoV-2 positive test until the end of follow-up. Nirmatrelvir group (N=9217) and control group (N=47,123). Adjusted hazard ratios and 95% confidence intervals are presented. Length of the bar represents the risk reduction per 100 persons at 90 days and associated 95% confidence intervals are also shown.

**Figure 2:**
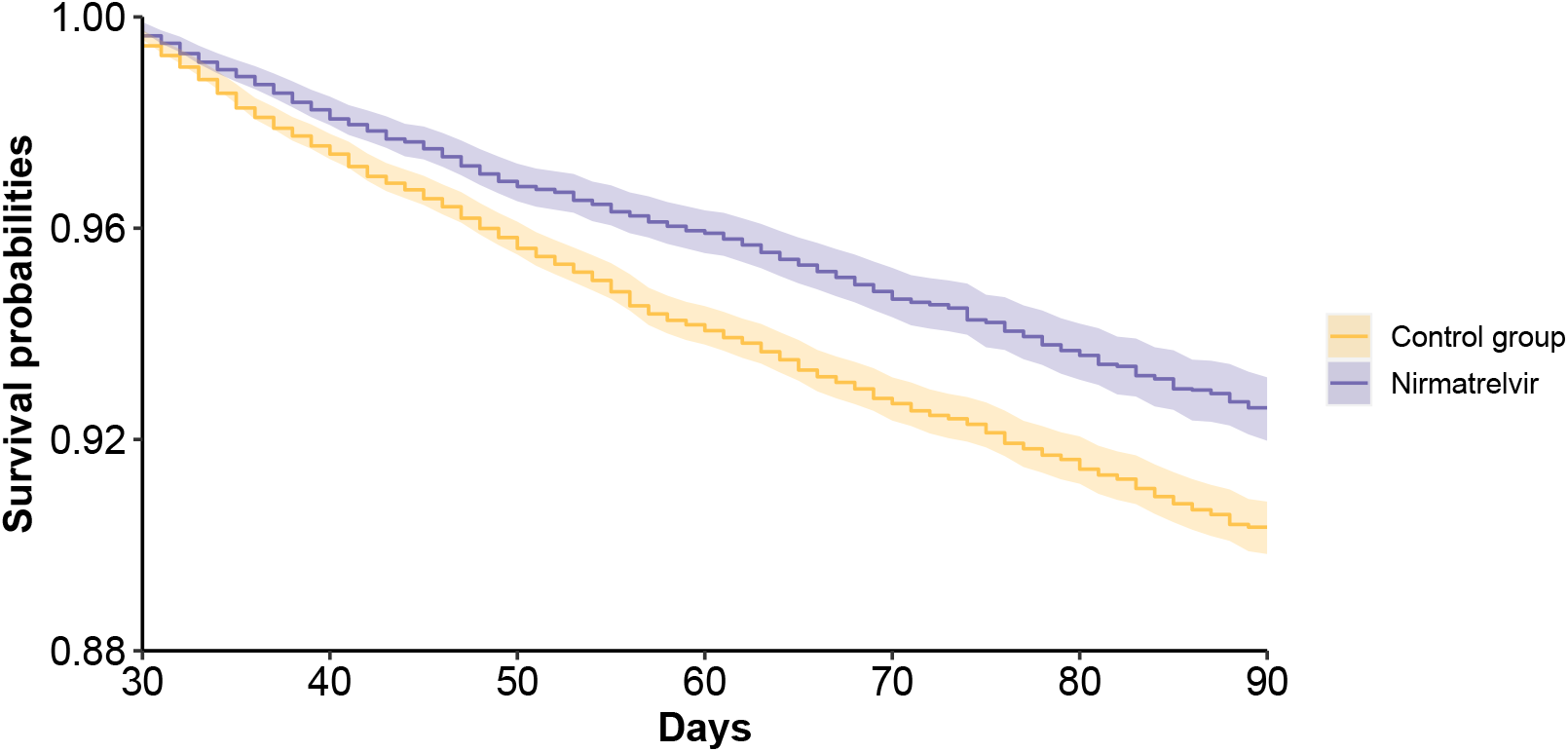
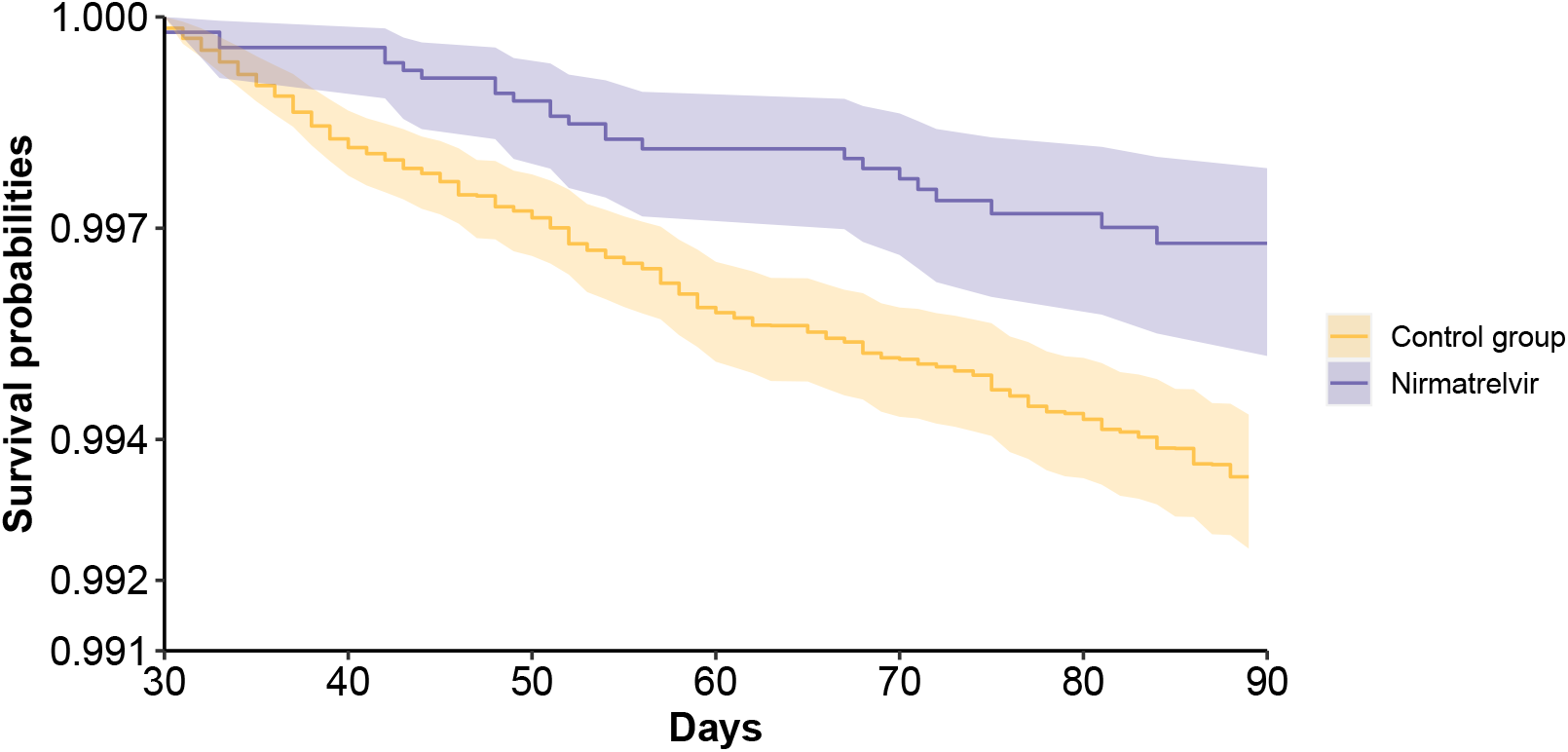
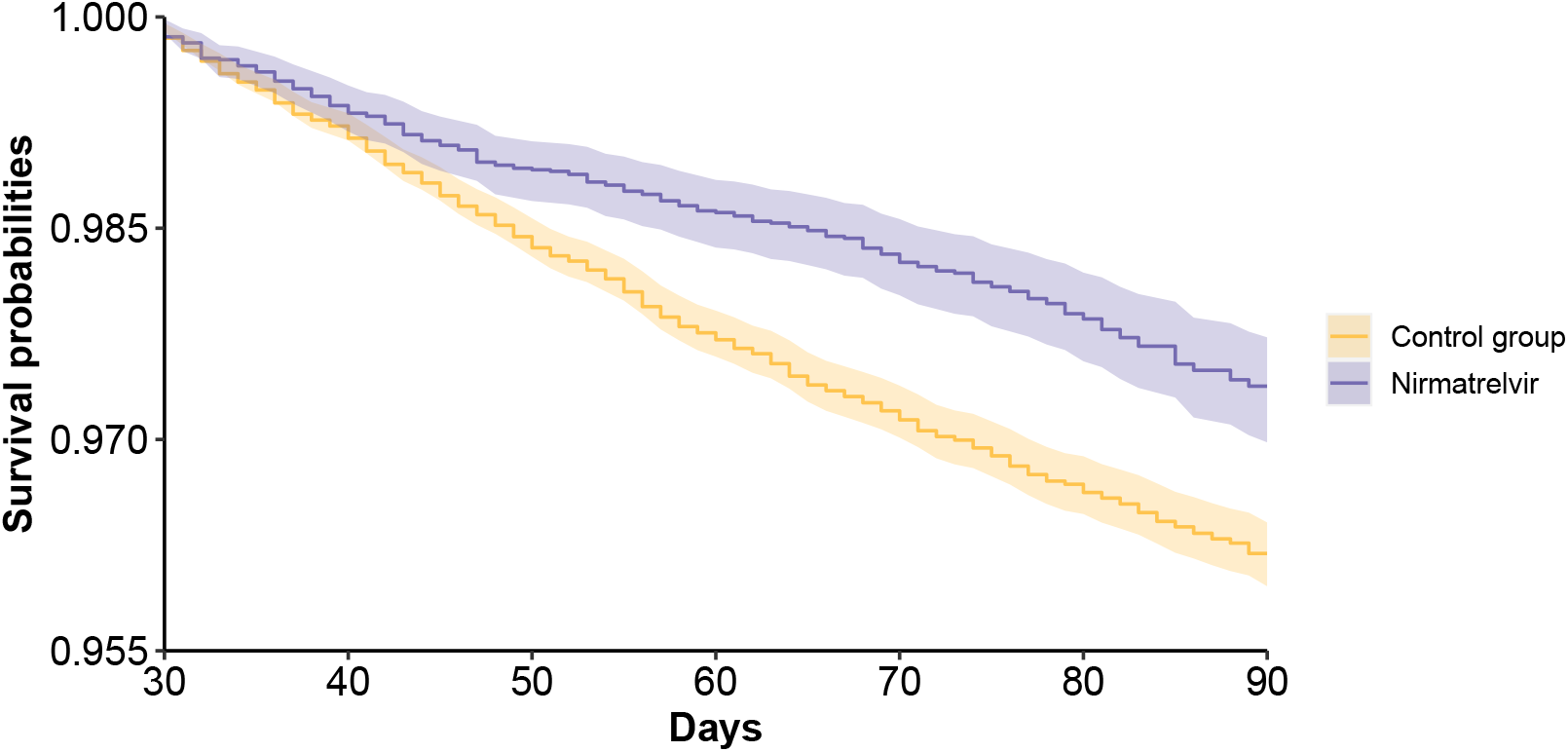
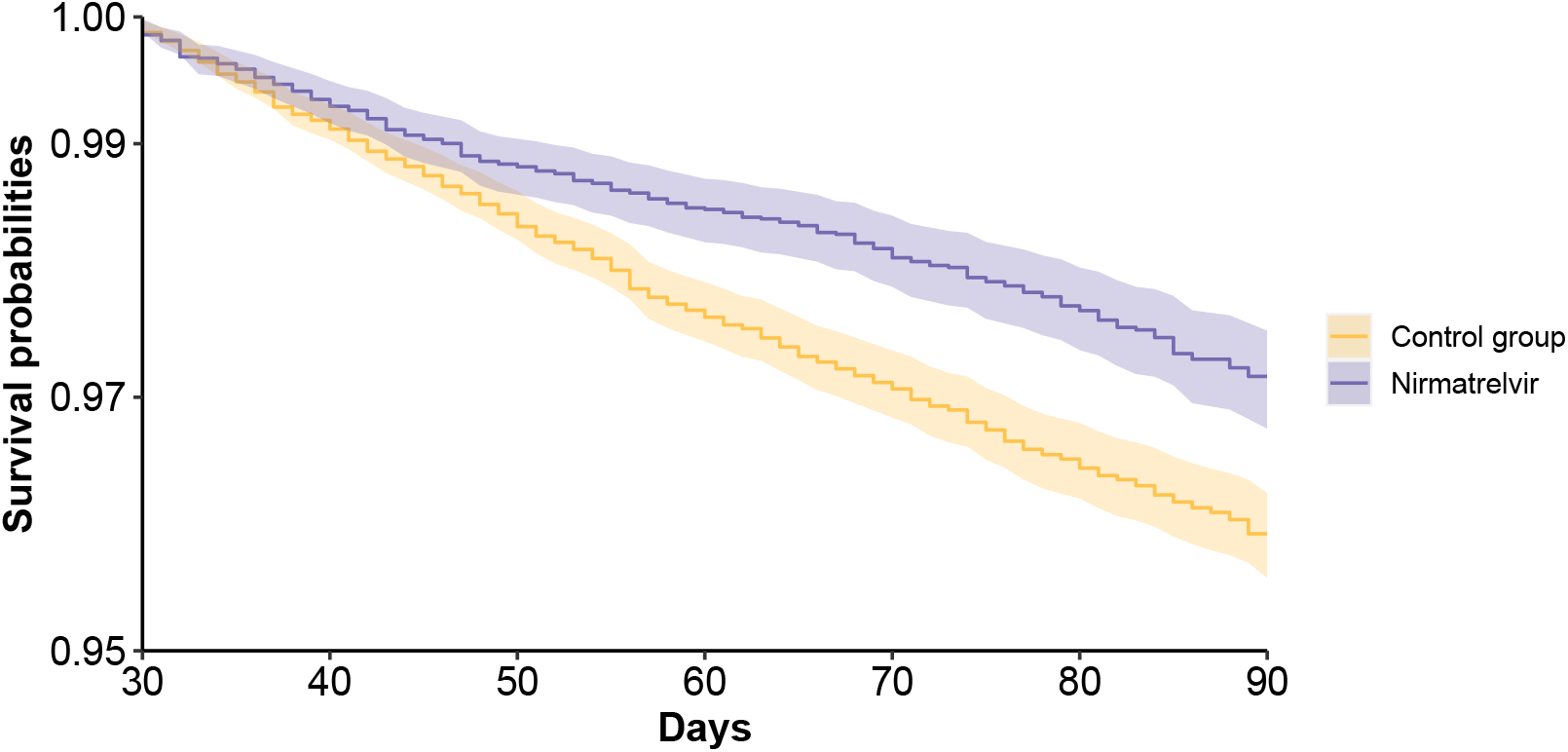
Survival probability of post-acute sequelae in nirmatrelvir and no treatment control group. a. post-acute sequelae of COVID-19 (PASC); b. death; c. hospitalization; d. composite outcome of death or hospitalization. Outcomes were ascertained 30 days after the SARS-CoV-2 positive test until the end of follow-up. Survival probability presented for nirmatrelvir (purple, N=9217) and control group (orange, N=47,123). Shaded areas are 95% confidence intervals.

### Risk of post-acute death and hospitalization

Compared to the control group, nirmatrelvir was associated with reduced risk of post-acute death (HR 0.52 (0.35, 0.77); ARR 0.28 (0.14, 0.41)), hospitalization (HR 0.70 (0.61, 0.80); ARR 1.09 (0.72, 1.46)), and the composite outcome of death or hospitalization (HR 0.67 (0.59, 0.77); ARR 1.32 (0.93, 1.70) (Figure 1 and 2b-d, supplemental table 2).

### Risk of individual post-acute sequela

Compared to the control group, nirmatrelvir was associated with reduced risk of 10 of the 12 pre-specified post-acute sequelae evaluated in this analysis.

Nirmatrelvir was associated with reduced risk of sequelae in the cardiovascular system (dysrhythmia and ischemic heart disease), coagulation and hematologic disorders (deep vein thrombosis, and pulmonary embolism), fatigue, liver disease, acute kidney disease, muscle pain, neurocognitive impairment, and shortness of breath (Figure 3, supplemental table 3).

**Figure 3:**
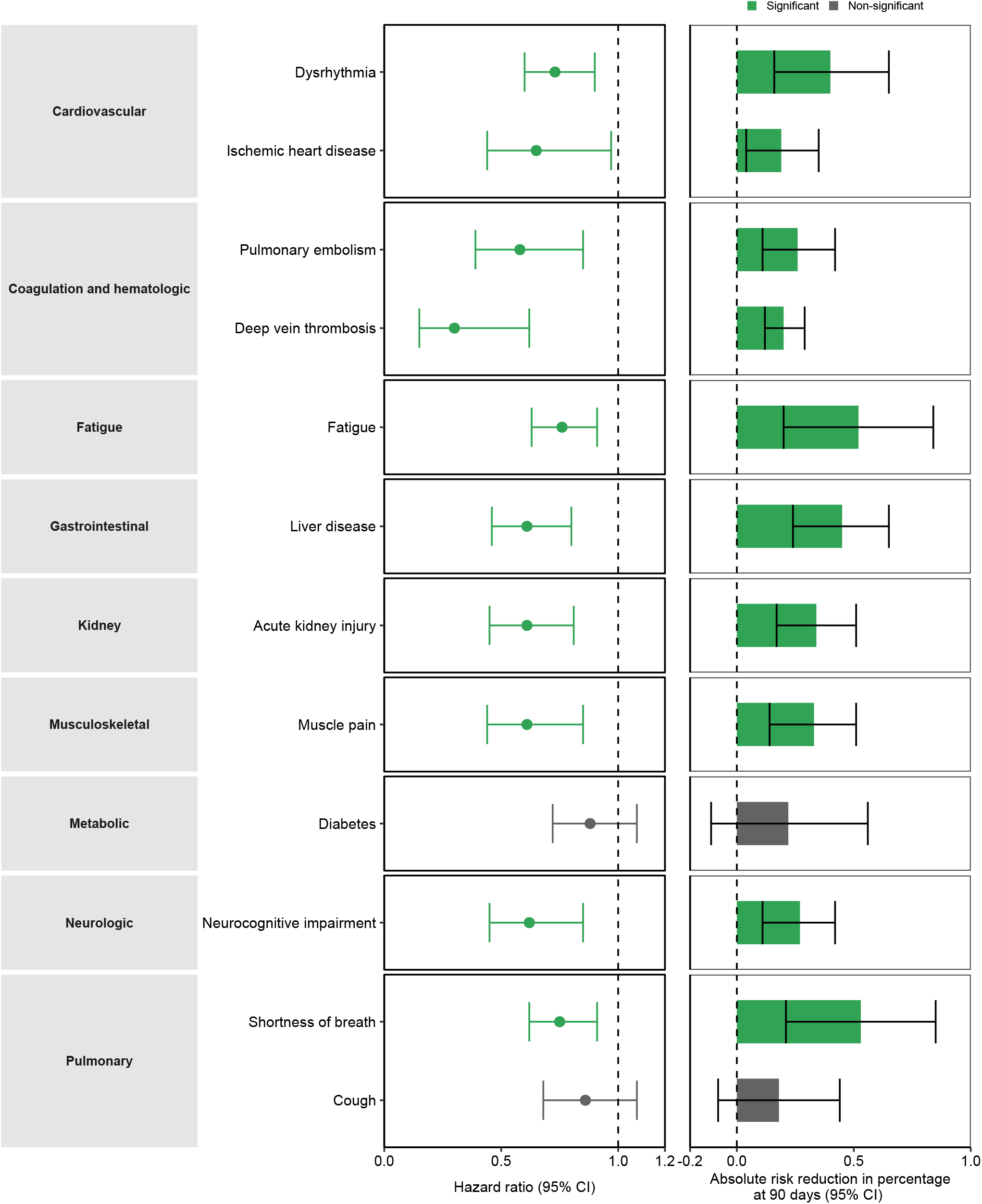
Hazard ratio and absolute risk reduction of nirmatrelvir on individual post-acute sequelae compared to no treatment control group. Outcomes were ascertained 30 days after the COVID-19 positive test until the end of follow-up. Nirmatrelvir group (N=9217) and control group (N=47,123). Adjusted hazard ratios and 95% confidence intervals are presented. Length of the bar represents the risk reduction per 100 persons at 90 days and associated 95% confidence intervals are also shown. Statistically significant results presented in green and results lack of statistical significance presented in gray.

There was lack of statistically significant association between nirmatrelvir and post-acute sequelae including new onset diabetes and cough (Figure 3, supplemental table 3).

### Risk of PASC in subgroups

Compared to the control group, people treated with nirmatrelvir exhibited reduced risk of PASC in subgroups based on age, race, sex, smoking, cancer, cardiovascular disease, chronic kidney disease, chronic lung disease, diabetes, immune dysfunction and hypertension (supplemental table 4).

Because nirmatrelvir is prescribed to people with at least one baseline risk factor for progression to severe acute COVID-19 illness, and to better understand the association between nirmatrelvir and the risk of PASC in people with different risk strata, we tested the association between nirmatrelvir and the risk of PASC according to the number of baseline risk factors for progression to severe acute COVID-19 illness. Nirmatrelvir was associated with reduced risk of PASC in people with 1 to 2, 3 to 4, and 5 or more baseline risk factors (supplemental table 4).

Examination of the association between nirmatrelvir and risk of PASC by vaccine status suggested that nirmatrelvir was associated with reduced risk of PASC in people who were unvaccinated, vaccinated, and those who received a booster vaccine (supplemental table 4).

Nirmatrelvir was associated with reduced risk of PASC in people with primary SARS-CoV-2 infection and in people with reinfection (supplemental table 4).

### Sensitivity analyses

To assess the robustness of our findings, we conducted multiple sensitivity analyses. 1) We applied the overlap weighting method to balance baseline characteristics in the treatment and control groups instead of the inverse probability weighing approach used in the primary analyses; 2) We applied the doubly robust approach to additionally adjust for covariates in the inverse probability weighted survival model, compared to the primary approach which used inverse probability weighted survival models; 3) We used a high dimensional variable selection algorithm to identify additional 100 covariates which were then used to along with a predefined set of covariates to construct the weights, compared to the primary approach which used predefined covariates; 4) We adjusted for health care utilization during follow up (including number of outpatient encounters, hospitalizations, laboratory tests and medications) as time varying covariates, compared to the primary approach which used covariate information at T_0_. All sensitivity analyses yielded results that are consistent (in both direction and magnitude) to those obtained using the primary approach (supplemental table 5).

## Discussion

In this study of 56,340 people with SARS-CoV-2 infection who had at least 1 risk factor for progression to severe COVID-19 illness, compared to the control group of people who did not receive antiviral or antibody treatment during the acute phase of SARS-CoV-2 infection, treatment with nirmatrelvir within 5 days of a positive SARS-CoV-2 test was associated with reduced risk of PASC including reduced risk of 10 of 12 post-acute sequelae examined.

Nirmatrelvir was also associated with reduced risk of post-acute death and hospitalization at 90 days. Nirmatrelvir was associated with reduced risk of PASC in subgroups based on age, race, sex, smoking, cancer, cardiovascular disease, chronic kidney disease, chronic lung disease, diabetes, immune dysfunction, and hypertension. Nirmatrelvir was associated with reduced risk of PASC across strata of baseline risk, and in people who were unvaccinated, vaccinated, and boosted; and in people with primary SARS-CoV-2 infection and reinfection. Altogether, the findings suggest that treatment with nirmatrelvir during the acute phase reduces the risk of post-acute adverse health outcomes.

Our results show that the salutary effect of nirmatrelvir extends to the post-acute phase of COVID-19; nirmatrelvir was associated with reduced risk of PASC in the overall cohort and in various subgroups including those across risk strata, vaccination status and regardless prior history of COVID-19. These findings are coupled with the observation that among 56,340 people with acute SARS-CoV-2 infection who had at least 1 risk factor for progression to severe disease who would be eligible for treatment with nirmatrelvir, 9217 (16.40%) were treated with nirmatrelvir and 47,123 (83.60%) received no antiviral treatment. The totality of evidence suggests the need to improve uptake and utilization of nirmatrelvir in the acute phase as a means of not only preventing progression to severe acute disease, but to also reduce the risk of post-acute adverse health outcomes.

Nirmatrelvir was associated with 26% less risk of PASC, 48% less risk of post-acute death, and 30% less risk of post-acute hospitalization; the magnitude of risk reduction on the absolute scale is also substantial amounting to 2.32, 0.28, and 1.09 less cases of PASC, post-acute death, and post-acute hospitalization for every 100 treated persons between 30 to 90 days of infection. Although the magnitude of risk reduction of post-acute outcomes reported here is more modest than the reported effect during the acute phase (5 to 6% less hospitalization or death per 100 treated persons during the first 30 days of infection), the clinical decision to initiate treatment with nirmatrelvir should consider its overall effectiveness in reducing burden of death and disease in both the acute and post-acute phase of COVID-19.

Nirmatrelvir was approved in the US for the treatment of acute COVID-19 illness in people with one or more risk factors for progression to severe disease. Our results evaluating the risk of PASC according to number of baseline risk factors suggest that the benefit was evident in people who had 1 to 2 baseline risk factors, progressively increased in a graded fashion as number of risk factors increased, and was most pronounced in people with 5 or more risk factors. These analyses suggest that those who are at most risk will likely derive the most benefit. Whether the salutary benefit of nirmatrelvir extends to people without risk factors for progression to severe disease (who would not qualify for nirmatrelvir prescription under the current FDA emergency use authorization and were not included in our analyses) remains to be tested in future randomized trials.

We note that our results suggested risk reduction for some but not all the pre-specified post-acute sequela in this analysis. It is possible that various sequelae are mediated by various mechanisms including some that may be affected by the receipt of antivirals and others that may not. Participants in our study were treated in the acute phase with a 5-day course of nirmatrelvir; it remains unclear whether longer duration or higher dose or both may have resulted in more reduced risk of post-acute sequelae. It is also unclear whether initiation of treatment in the post-acute phase of COVID-19 reduces the risk of Long Covid.

This study has several strengths. The VA operates the largest integrated health care system in the US, and the vast and rich national healthcare databases of the US Department of Veterans Affairs–with larger number of treated and untreated patients followed longitudinally over time allows the evaluation of outcomes that were not assessed in randomized trials. VA data contains comprehensive information about participants including COVID-19 testing, medication use including vaccination records, hospitalization records, death records, and other attributes, which allows the comprehensive capture of covariates from different data domains including demographics, diagnoses, laboratory test results, medications, vital signs, health care utilization and contextual factors. We tested robustness of our findings in multiple sensitivity analyses which yielded consistent results.

This study has several limitations. The demographic composition of our cohort (majority White and male) may limit generalizability of study findings. We used the electronic healthcare databases of the US Department of Veterans Affairs to conduct this study, and although we took care to adjust the analyses for a large set of predefined variables, we cannot completely rule out misclassification bias and residual confounding. We did not capture nirmatrelvir use outside the VA system; if large number of people in the control group used nirmatrelvir outside the VA, this may bias the results toward the null. We focused our analyses on a pre-specified set of 12 sequelae and did not examine all sequelae of COVID-19. Finally, as the virus continues to mutate and as new variants emerge and as vaccine uptake improves, it is possible that the real-world effectiveness of nirmatrelvir may also change over time.

In sum, we provide evidence in this study that in people with SARS-CoV-2 infection who had at least 1 risk factor for progression to severe disease, treatment with nirmatrelvir within 5 days of a positive SARS-CoV-2 test was associated with reduced risk of PASC across the risk spectrum in this cohort and regardless of vaccination status and history of prior infection. The findings suggest that the salutary benefit of nirmatrelvir extends to the post-acute phase of COVID-19.

## Data Availability

Data is available from the US Department of Veterans Affairs

## Acknowledgements

This study used data from the VA COVID-19 Shared Data Resource. This research was funded by the United States Department of Veterans Affairs (ZAA). The funders had no role in study design, data collection and analysis, decision to publish or preparation of the manuscript. The contents do not represent the views of the US Department of Veterans Affairs or the US government.

## Author Contributions

YX and ZAA contributed to the development of the study concept and design. YX, TC, and ZAA contributed to data analysis. YX and ZAA contributed to interpretation of the results. YX and ZAA drafted the manuscript. Critical revision of the manuscript was contributed to by YX and ZAA. YX, TC, and ZAA developed the data visualization. ZAA provided administrative, technical, and material support. ZAA provided supervision and mentorship. ZAA is the guarantor of the work. Each author contributed important intellectual content during manuscript drafting or revision and accepts accountability for the overall work by ensuring that questions pertaining to the accuracy or integrity of any portion of the work are appropriately investigated and resolved. All authors approved the final version of the report. The corresponding author attests that all the listed authors meet the authorship criteria and that no others meeting the criteria have been omitted.

## Competing interests

The authors declare no conflict of interest.

## Funding/Support

This research was funded by the United States Department of Veterans Affairs (for ZAA).

## Disclaimer

The contents do not represent the views of the US Department of Veterans Affairs or the US government.

## Figure Legends

**Figure 4: Hazard ratio and absolute risk reduction of nirmatrelvir on post-acute sequelae of COVID-19 compared to no treatment control group by subgroups.** a. by demographic and disease subgroups included age (≤60, >60-≤70 and >70), race (White and Black), sex, smoking status (current smoker, former smoker and never smoker), cancer, cardiovascular disease, chronic kidney disease, chronic lung disease, diabetes, immune dysfunction, and hypertension.; b. by number of baseline risk factors (1-2, 3-4, ≥5), vaccination status (unvaccinated, 1-2 doses of vaccine and boosted), and SARS-CoV2 infection status (with primary SARS-CoV-2 infection and reinfection). Baseline risk factors of progression to severe acute COVID-19 illness included age>60, BMI>25 km/m^2^, current smoker, cancer, cardiovascular disease, kidney disease, chronic lung disease, diabetes, immune dysfunction and hypertension. Outcomes were ascertained 30 days after the SARS-CoV-2 positive test until the end of follow-up. Plot represents the risk reduction per 100 persons at 90 days and associated 95% confidence intervals.

## Notes

### Competing Interest Statement

The authors have declared no competing interest.

### Author Declarations

The Institutional Review Board of the VA Saint Louis Health Care System gave approval of this study.

