## Supplemental for "Nirmatrelvir and the Risk of Post-Acute Sequelae of COVID-19"

**Supplemental Figure 1. Cohort flow**

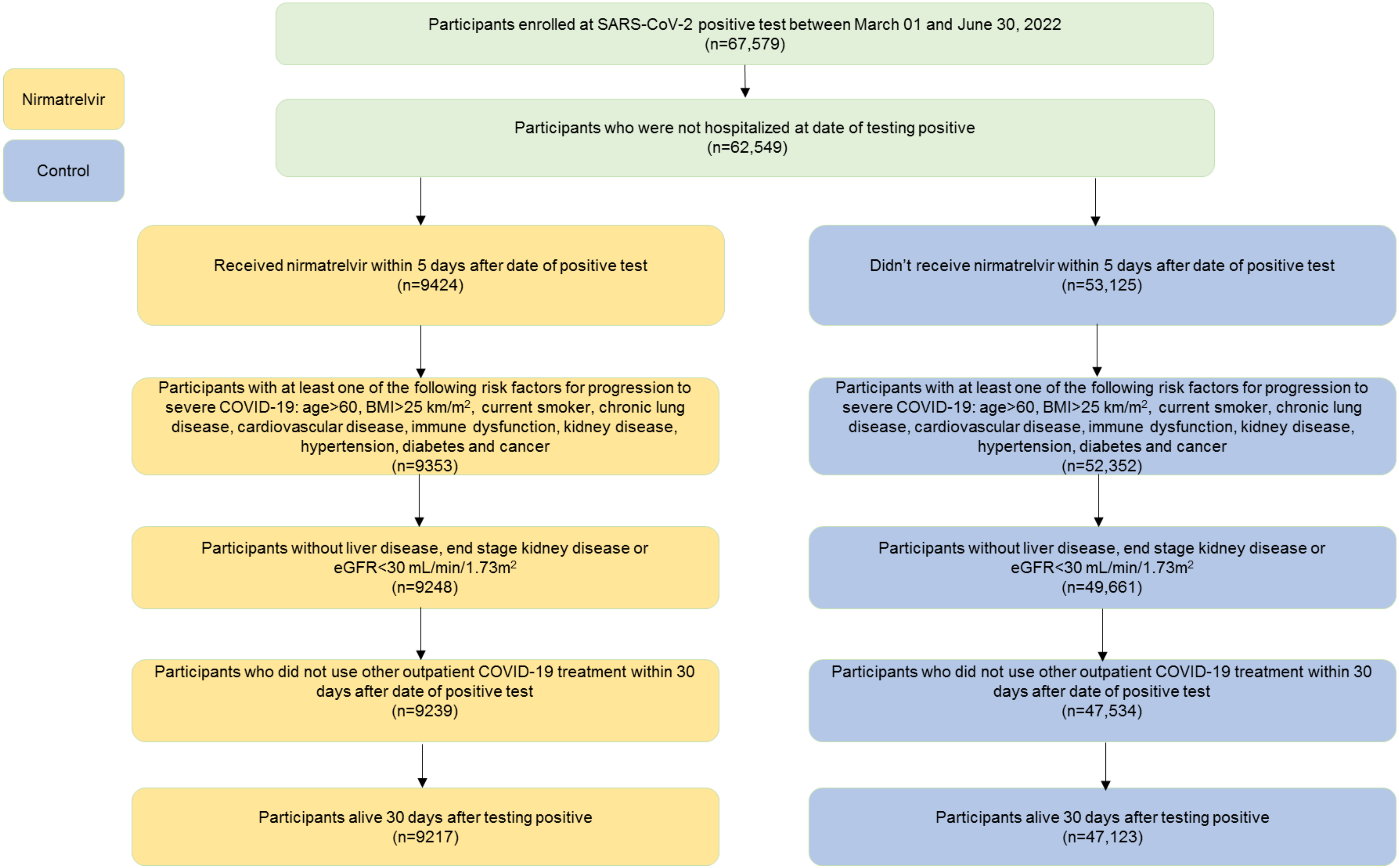

**Supplemental Figure 2. Cohort timeline**

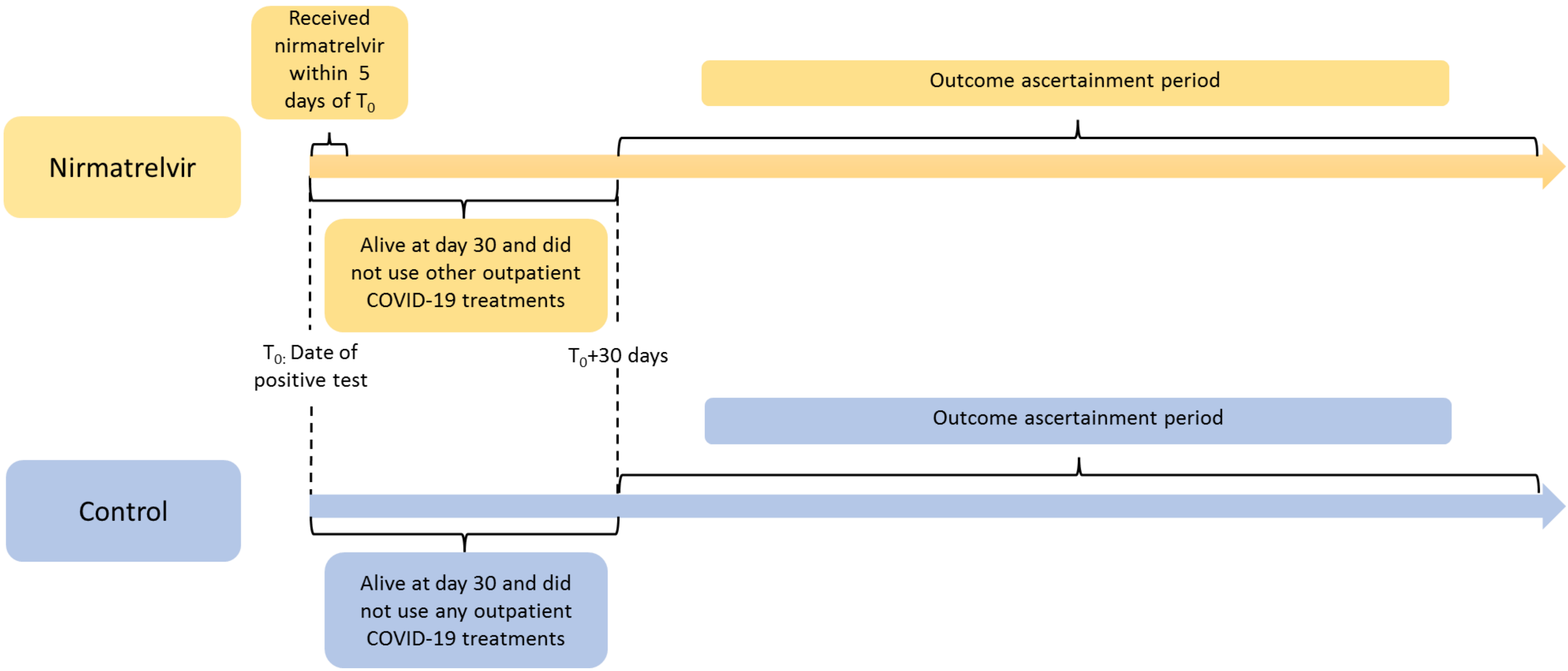

**Supplemental Table 1. Demographic and health characteristics of the overall cohort, the nirmatrelvir group, and the control group before weighting**

| | Overall cohort<br>N = 56,340 | Nirmatrelvir group<br>N = 9217 | Control group<br>N = 47,123 | SMD between Nirmatrelvir and control group |
|---|---|---|---|---|
| **Age, mean (std), yr** | 62.85 (14.91) | 65.06 (13.92) | 62.41 (15.05) | 0.18 |
| **Race, no. (%)** | | | | |
| White | 40,602 (72.07) | 6873 (74.57) | 33,729 (71.58) | 0.07 |
| Black | 11,638 (20.66) | 1757 (19.06) | 9881 (20.97) | 0.05 |
| Other | 4100 (7.28) | 587 (6.37) | 3513 (7.46) | 0.04 |
| **Sex, no. (%)** | | | | |
| Male | 48,301 (85.73) | 8072 (87.58) | 40,229 (85.37) | 0.07 |
| Female | 8039 (14.27) | 1145 (12.42) | 6894 (14.63) | 0.07 |
| **Smoking status, no. (%)** | | | | |
| Never | 26,537 (47.10) | 4094 (44.42) | 22,443 (47.63) | 0.06 |
| Former | 20,412 (36.23) | 3859 (41.87) | 16,553 (35.13) | 0.14 |
| Current | 9391 (16.67) | 1264 (13.71) | 8127 (17.25) | 0.10 |
| **Area Deprivation Index*, mean (std)** | 50.73 (19.98) | 48.31 (20.01) | 51.2 (19.94) | 0.14 |
| **Vaccination, no. (%)** | | | | |
| **Without prior vaccination** | 14,521 (25.77) | 1510 (16.38) | 13,011 (27.61) | 0.27 |
| **With 1 shots of vaccination** | 2504 (4.44) | 334 (3.62) | 2170 (4.61) | 0.05 |
| **With 2 shots of vaccination** | 13,980 (24.81) | 2170 (23.54) | 11,810 (25.06) | 0.04 |
| **With vaccinate booster** | 25,335 (44.97) | 5203 (56.45) | 20,132 (42.72) | 0.28 |
| **BMI, mean (std)** | 30.61 (5.22) | 30.94 (5.24) | 30.55 (5.21) | 0.07 |
| **eGFR, mean (std), ml/min/1.73m$^2$** | 79.97 (19.32) | 78.76 (18.50) | 80.21 (19.47) | 0.08 |
| **Systolic blood pressure, mean (std), mmHg** | 132.60 (11.65) | 133.64 (11.22) | 132.39 (11.73) | 0.11 |
| **Diastolic blood pressure, mean (std), mmHg** | 78.26 (7.09) | 78.21 (6.94) | 78.27 (7.11) | 0.01 |
| **History of SARS-CoV-2 infection, no. (%)** | 4471 (7.94) | 704 (7.64) | 3767 (7.99) | 0.01 |
| **Use of steroid, no. (%)** | 5854 (10.39) | 813 (8.82) | 5041 (10.70) | 0.06 |
| **Cancer, no. (%)** | 7476 (13.27) | 1385 (15.03) | 6091 (12.93) | 0.06 |
| **Chronic lung disease, no. (%)** | 12,256 (21.75) | 2111 (22.90) | 10,145 (21.53) | 0.03 |
| **Dementia, no. (%)** | 4666 (8.28) | 743 (8.06) | 3923 (8.33) | 0.01 |
| **Diabetes mellitus type 2, no. (%)** | 17,176 (30.49) | 3128 (33.94) | 14,048 (29.81) | 0.09 |
| **Cardiovascular disease, no. (%)** | 16,569 (29.41) | 2848 (30.90) | 13,721 (29.12) | 0.04 |

| | | | | |
|---|---|---|---|---|
| **Hyperlipidemia, no. (%)** | 41,114 (72.97) | 7461 (80.95) | 33,653 (71.42) | 0.23 |
| **Immune dysfunction, no. (%)** | 2542 (4.51) | 534 (5.79) | 2008 (4.26) | 0.07 |
| **Hospital bed capacity†, mean (std)** | 477.33 (296.37) | 475.88 (303.17) | 477.61 (295.03) | 0.01 |
| **Hospital bed occupancy†, mean (std)** | 0.43 (0.13) | 0.43 (0.13) | 0.43 (0.13) | 0.03 |
| **Number of hospital admissions‡, mean (std)** | 0.20 (0.79) | 0.17 (0.66) | 0.21 (0.81) | 0.05 |
| **Number of outpatient visits‡, mean (std)** | 2.81 (1.58) | 3.20 (1.39) | 2.74 (1.61) | 0.31 |
| **Number of blood panel tests‡, mean (std)** | 7.15 (9.26) | 7.54 (7.82) | 7.07 (9.51) | 0.05 |
| **Number of medications‡, mean (std)** | 8.90 (7.70) | 10.00 (7.18) | 8.68 (7.78) | 0.18 |
| **Influenza vaccine, no. (%)** | 35,000 (62.12) | 6627 (71.90) | 28,373 (60.21) | 0.25 |
| **Calendar week of study enrollment, mean (std)** | 19.99 (4.67) | 21.23 (3.59) | 19.75 (4.81) | 0.35 |
| **Follow up, days IQR** | 93 (73, 118) | 86 (70, 106) | 94 (73, 121) | |

*. Area Deprivation Index is a measure of socioeconomic disadvantage, with a range from low to high disadvantage of 0 to 100.
†. Data collected as average value during the week of positive SARS-CoV-2 test result in the hospital where the test was performed.
‡. Data collected within one year before the study enrollment.
BMI, body mass index; eGFR, estimated glomerular filtration rate; IQR, interquartile range; SMD, absolute standardized mean difference; std, standard deviation.

**Supplemental Table 2. Hazard ratio and absolute risk reduction of nirmatrelvir on post-acute sequelae of COVID-19, death, hospitalization, and composite outcome of death or hospitalization compared to control group**

| Outcome | Hazard Ratio | Nirmatrelvir group event rate in % at 90 days (95% CI) | Control group event rate in % at 90 days (95% CI) | Absolute risk reduction in % at 90 days |
|---|---|---|---|---|
| **Post-acute sequelae of COVID-19** | 0.74 (0.69, 0.81) | 7.11 (6.57, 7.64) | 9.43 (9.14, 9.72) | 2.32 (1.73, 2.91) |
| **Post-acute death** | 0.52 (0.35, 0.77) | 0.30 (0.19, 0.42) | 0.58 (0.50, 0.66) | 0.28 (0.14, 0.41) |
| **Post- acute hospitalization** | 0.70 (0.61, 0.80) | 2.57 (2.23, 2.90) | 3.66 (3.46, 3.85) | 1.09 (0.72, 1.46) |
| **Post-acute death or hospitalization** | 0.67 (0.59, 0.77) | 2.78 (2.43, 3.12) | 4.09 (3.89, 4.29) | 1.32 (0.93, 1.70) |

Nirmatrelvir group defined as received nirmatrelvir within 5 days after tested positive for SARS-CoV-2 and did not use any other outpatient antiviral or antibody during first 30 days after tested positive.
Control group defined as did not use any outpatient antiviral or antibody during first 30 days after tested positive and served as reference group in the analyses.
Outcomes were ascertained 30 days after the SARS-CoV-2 positive test until the end of follow-up.
CI, confidence interval

**Supplemental Table 3. Hazard ratio and absolute risk reduction of nirmatrelvir on individual post-acute sequelae compared to control group**

| Organ system | Outcome | Hazard Ratio (95% CI) | Nirmatrelvir group event rate in % at 90 days (95% CI) | Control group event rate in % at 90 days (95% CI) | Absolute risk reduction % at 90 days (95% CI) |
|---|---|---|---|---|---|
| **Cardiovascular** | **Dysrhythmia** | 0.73 (0.60, 0.90) | 1.12 (0.91, 1.34) | 1.53 (1.40, 1.65) | 0.40 (0.16, 0.65) |
| | **Ischemic heart disease** | 0.65 (0.44, 0.97) | 0.36 (0.23, 0.50) | 0.56 (0.48, 0.63) | 0.19 (0.04, 0.35) |
| **Coagulation and hematologic** | **Deep vein thrombosis** | 0.30 (0.15, 0.62) | 0.09 (0.03, 0.15) | 0.29 (0.24, 0.35) | 0.20 (0.12, 0.29) |
| | **Pulmonary embolism** | 0.58 (0.39, 0.85) | 0.36 (0.23, 0.49) | 0.62 (0.54, 0.70) | 0.26 (0.11, 0.42) |
| **Fatigue** | **Fatigue** | 0.76 (0.63, 0.91) | 1.64 (1.35, 1.93) | 2.17 (2.01, 2.32) | 0.52 (0.20, 0.84) |
| **Gastrointestinal** | **Liver disease** | 0.61 (0.46, 0.80) | 0.69 (0.51, 0.87) | 1.14 (1.03, 1.25) | 0.45 (0.24, 0.65) |
| **Kidney** | **Acute kidney injury** | 0.61 (0.45, 0.81) | 0.53 (0.38, 0.68) | 0.88 (0.78, 0.97) | 0.34 (0.17, 0.51) |
| **Musculoskeletal** | **Muscle pain** | 0.61 (0.44, 0.85) | 0.52 (0.36, 0.69) | 0.85 (0.76, 0.95) | 0.33 (0.14, 0.51) |
| **Metabolic** | **Diabetes** | 0.88 (0.72, 1.08) | 1.62 (1.31, 1.93) | 1.85 (1.69, 2.00) | 0.22 (-0.11, 0.56) |
| **Neurological** | **Neurocognitive impairment** | 0.62 (0.45, 0.85) | 0.44 (0.30, 0.57) | 0.70 (0.62, 0.78) | 0.27 (0.11, 0.42) |
| **Pulmonary** | **Shortness of breath** | 0.75 (0.62, 0.91) | 1.65 (1.35, 1.94) | 2.18 (2.02, 2.33) | 0.53 (0.21, 0.85) |
| | **Cough** | 0.86 (0.68, 1.08) | 1.09 (0.85, 1.32) | 1.27 (1.15, 1.39) | 0.18 (-0.08, 0.44) |

Nirmatrelvir group defined as received nirmatrelvir within 5 days after tested positive for SARS-CoV-2 and did not use any other outpatient antiviral or antibody during first 30 days after tested positive.
Control group defined as did not use any outpatient antiviral or antibody during first 30 days after tested positive and served as reference group in the analyses.
Outcomes were ascertained 30 days after the SARS-CoV-2 positive test until the end of follow-up.
CI, confidence interval

**Supplemental Table 4. Hazard ratio of post-acute sequelae of COVID-19 in nirmatrelvir compared to control by subgroups**

| Subgroup | Description | Hazard Ratio (95% CI) |
|---|---|---|
| Age | Age $\leq$ 60 | 0.72 (0.62, 0.83) |
| | 60 $<$ Age $\leq$ 70 | 0.81 (0.70, 0.92) |
| | Age $>$ 70 | 0.74 (0.68, 0.82) |
| Race | White | 0.73 (0.67, 0.79) |
| | Black | 0.85 (0.73, 0.99) |
| Sex | Male | 0.75 (0.70, 0.81) |
| | Female | 0.73 (0.58, 0.91) |
| Smoking status | Current | 0.71 (0.59, 0.85) |
| | Former | 0.76 (0.69, 0.84) |
| | Never | 0.75 (0.68, 0.84) |
| Cancer | No | 0.74 (0.68, 0.80) |
| | Yes | 0.77 (0.67, 0.89) |
| Cardiovascular disease | No | 0.74 (0.67, 0.81) |
| | Yes | 0.76 (0.69, 0.83) |
| Chronic kidney disease | No | 0.74 (0.68, 0.80) |
| | Yes | 0.77 (0.67, 0.89) |
| Chronic lung disease | No | 0.76 (0.70, 0.82) |
| | Yes | 0.72 (0.64, 0.82) |
| Diabetes | No | 0.73 (0.67, 0.80) |
| | Yes | 0.77 (0.70, 0.86) |
| Immune dysfunction | No | 0.74 (0.69, 0.80) |
| | Yes | 0.78 (0.61, 1.01) |
| Hypertension | No | 0.80 (0.74, 0.86) |

| | | |
|---|---|---|
| | **Yes** | 0.65<br>(0.57, 0.74) |
| **Number of baseline risk factors*** | ≤ **2** | 0.77<br>(0.67, 0.88) |
| | **3** or **4** | 0.77<br>(0.72, 0.83) |
| | ≥ **5** | 0.70<br>(0.61, 0.80) |
| **Vaccine** | **Unvaccinated** | 0.68<br>(0.57, 0.82) |
| | **1 or 2 doses of vaccine** | 0.69<br>(0.64, 0.75) |
| | **Boosted** | 0.79<br>(0.72, 0.86) |
| **SARS-CoV-2 infection** | **Primary infection** | 0.75<br>(0.70, 0.81) |
| | **Reinfection** | 0.75<br>(0.66, 0.84) |

**Supplemental Table 5. Sensitivity analyses**

| Sensitivity analyses | Hazard Ratio for risk of PASC (95% CI) |
|---|---|
| **Balance through overlap weighting** | 0.74 (0.69, 0.80) |
| **Doubly robust adjustment** | 0.77 (0.72, 0.82) |
| **Additionally adjust for 100 high dimensional variables** | 0.75 (0.70, 0.80) |
| **Adjusting for time varying health care utilization** | 0.79 (0.73, 0.85) |

Hazard ratios for nirmatrelvir group compared to control group on the risk of post-acute sequelae of COVID-19 were reported.
CI, confidence interval
